# Impacts of the Pacific Northwest Heat Dome on preterm birth rates in Oregon and Washington State

**DOI:** 10.64898/2025.12.17.25342502

**Authors:** Heather McBrien, Ralph Catalano, Tim Bruckner, Nina M. Flores, Allison Stolte, Alison Gemmill, Joan A. Casey

## Abstract

Acute heat exposure, which is increasing with climate change, likely increases preterm birth risk. However, few studies consider susceptible exposure windows for extreme heat events, particularly among historically unexposed populations. The 2021 Pacific Northwest Heat Dome produced the highest temperatures ever recorded in usually temperate Oregon and Washington State, offering an ideal study setting. We used 2016–2022 vital statistics records to estimate the gestation month-specific impact of the Heat Dome on preterm birth. Using an interrupted time series design with a synthetic control, we compared the observed odds of preterm birth in the exposed (*in utero* November 2020–July 2021) Oregon and Washington conception cohorts to counterfactual odds had the Heat Dome not happened. Analysis included 716,096 exposed births across 67 monthly conception cohorts. We identified increased odds of preterm birth in cohorts exposed during months 2-3 (11% increase, 95% CI: [1%, 22%]) and 6-7 (14% increase, 95% CI: [5%, 24%]) of pregnancy. These findings partially agree with literature reporting elevated preterm birth risk after heat exposure in all trimesters. As extreme heat events are now expected once to twice per decade rather than once every thousand years, they pose risks to perinatal health.

## Introduction

The June 2021 Pacific Northwest Heat Dome (PNW Heat Dome) produced the highest ever recorded temperatures in Oregon and Washington State (48°C and 47°C, respectively)^1,2^ and led to 610 excess deaths.^3^ If climate change warms global temperatures 2°C above early-industrial levels, which nearly all climate scenarios predict,^5^ heatwaves like the PNW Heat Dome are expected once to twice per decade, compared to once every thousand years pre-climate change.^4^

Events like the PNW Heat Dome are especially dangerous in historically temperate places like Oregon and Washington, where buildings are designed to retain heat and often lack air conditioning.^6^ Because these places have not adapted to heat, during extreme heat, many people are exposed to unsafe temperatures and suffer health consequences like dehydration, heat sickness, and death.^7,8^

Extreme heat is especially dangerous for pregnant people. High-quality studies show short-term exposure to heat in the days or weeks prior to delivery increases risk of adverse outcomes, including preterm birth (<37 weeks gestation).^9,10,11^ However, few studies have quantified the risks of preterm birth after anomalous extreme heat emergencies, in historically temperate areas, or identified susceptible exposure windows.^9,10^

In this paper, we aimed to quantify how preterm birth risk changed in pregnancies exposed to the PNW Heat Dome in hardest-hit Oregon and Washington State. We used a quasi-experimental interrupted time-series method^12^ to determine the specific gestational months in which heat exposure increased preterm birth risk.^13^

## Methods

We used national vital statistics data on 716,096 live births in Oregon and Washington State conceived from January 1^st^, 2016 to July 31^st^, 2021. These records included birth month and year, best obstetric estimate of gestational age, and birthing person county of residence.^14^ We excluded births with gestational age < 20 weeks. We created 67 monthly conception cohorts from January 2016–July 2021 by aggregating monthly counts of all births to people residing in Oregon or Washington, and calculated monthly odds of preterm birth in each conception cohort (e.g. [number of births with gestational age <37 weeks conceived January 2016] / [number of births with gestational age ≥37 weeks conceived January 2016]). The PNW Heat Dome lasted from June 25^th^ to July 7^th^, 2021. We considered all conception cohorts from November 2020 to July 2021 exposed, hypothesizing that the Heat Dome increased odds of preterm birth in these conception cohorts.

Then, we created an analogous counterfactual time series of monthly odds of preterm birth in each “Oregon-Washington” conception cohort for the same period, meant to approximate the odds of preterm birth in these states had there been no Heat Dome. To build the counterfactual time series, we first constructed a synthetic control time series^15^ of monthly odds of preterm birth in each conception cohort for Oregon-Washington. The synthetic control time series was a weighted average of odds from unexposed counties which did not experience anomalous heat. These weights minimized the difference between observed pre-heatwave odds in affected Oregon-Washington and the synthetic control (i.e., unaffected counties) time series.

All Oregon and Washington counties were considered exposed. Counties outside of Oregon and Washington served as controls if county temperatures during the Heat Dome dates didn’t exceed the 90th percentile of historical temperatures on those dates 1981-2011. We built the synthetic control with counties rather than states because many states were partially exposed to extreme temperatures during the heatwave. Excluding all partially exposed states would have left insufficient data to approximate the preterm birth odds in Oregon and Washington.

November 2020 conception cohort pregnancies were exposed to the Heat Dome if they were *in utero* during gestational weeks 33-36, and all conception cohorts from October 2020 forward were exposed in earlier months of pregnancy. We fit a linear regression model to pre-Heat Dome observations (conception cohorts before November 2020), treating the synthetic control time series as the predictor and the observed odds time series as the outcome. We examined residuals from the regression using Box– Jenkins methods^16^ to detect autocorrelation, and added an ARIMA error to the regression to model this dependence, resulting in a Box–Jenkins transfer function. We used coefficients from this model to predict counterfactual odds of preterm birth had there been no Heat Dome in Oregon-Washington for the nine exposed conception cohorts November 2020 - July 2021.

We used a method by Chen and Liu^12^ to test for outliers in the differences between the observed and counterfactual odds of preterm birth in conception cohorts exposed to the Heat Dome. We tested for outlying sequences in the nine conception cohorts *in utero* during the Heat Dome, using an outlier-detection threshold of *t* = 1.96 (*p* < 0.05). Outliers indicated that the observed odds of preterm birth deviated more than expected from the counterfactual odds had the Heat Dome not happened, implying the Heat Dome affected the odds of preterm birth.

We conducted our analyses with Scientific Computing Associates’ time-series processor (http://www.scausa.com/scapc.php). The UC Irvine Committee for the Protection of Human Subjects approved this study (protocol # 20195444).

## Results

Our analysis included 716,096 exposed births in Oregon and Washington across 67 monthly conception cohorts from January 2016 to July 2021. The mean odds of preterm birth over the entire study period were 110 per 1000 pregnancies, ranging from 97 to 129 (**Figure 1A**). The mean odds of preterm birth in cohorts exposed *in utero* to the Heat Dome were 9.8% higher compared to earlier unexposed cohorts (118 per 1000 versus 108 per 1000).

**Figure 1.**
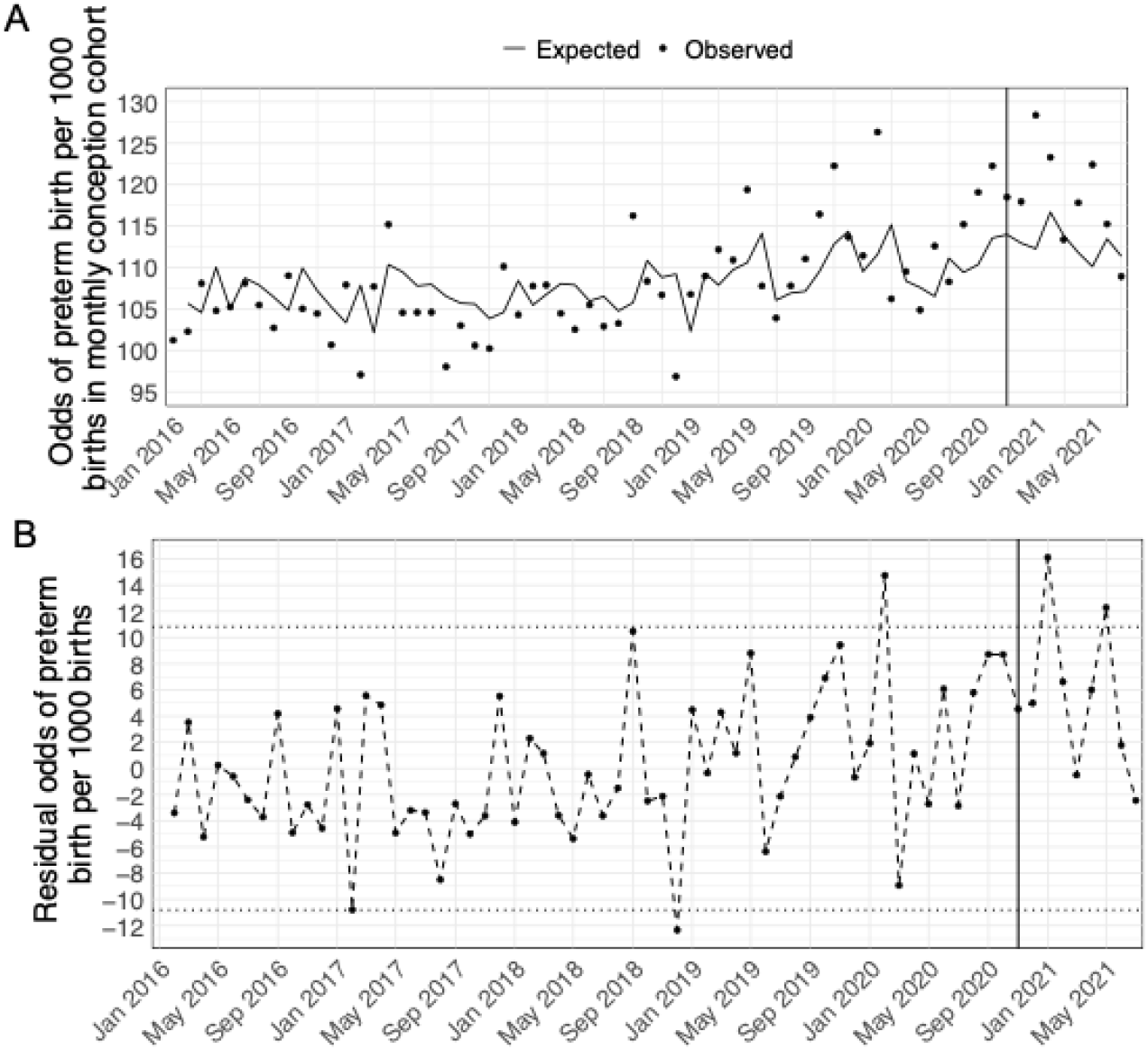
**<u>A: Observed and counterfactual (no PNW Heat Dome) odds of preterm birth in Oregon and Washington State conception cohorts January 2016 to July 2021</u>**. Figure 1A plots the odds of preterm birth per 1000 live births for conception cohorts in Oregon and Washington State January 2016 – July 2021. Points plot the observed odds of preterm birth (e.g. ({number of births with gestational age <37 weeks conceived January 2016} / {number of births with gestational age ≥37 conceived January 2016})* 1000), while the line plots the predicted counterfactual odds of preterm birth had the Pacific Northwest Heat Dome not happened, generated from a Box-Jenkins transfer function created with pre-heatwave data, with predictions carried through the heatwave. The vertical line indicates the first conception cohorts exposed *in utero* to the heatwave (November 2020). All counties in Oregon and Washington were considered exposed. Counties outside of Oregon and Washington served as controls for the synthetic control time series if temperatures didn’t exceed the 90th percentile of historical temperatures on Heat Dome days 1981-2011. Control counties were included from the following states: AL, AZ, AR, CA, CO, DE, FL, GA, ID, IL, IN, IA, KS, KY, LA, MD, MI, MN, MS, MO, MT, NE, NV, NJ, NM, NC, ND, OH, OK, PA, SC, SD, TN, TX, UT, VA, WV, WI, WY. **<u>B: Differences between observed and counterfactual (no PNW Heat Dome) odds of preterm birth in Oregon and Washington State conception cohorts January 2016 to July 2021</u>**. Figure 1B plots the residual incidence of preterm birth for Oregon and Washington conception cohorts after subtracting predicted odds of preterm birth from the Box-Jenkins transfer function created with pre-heatwave data from actual observed odds of preterm birth. The dotted lines represent 95% confidence intervals for outlier detection, and the vertical line indicates the first conception cohort exposed *in utero* to the heatwave (November 2020).

In the time series of differences between observed and expected odds of preterm birth in exposed cohorts (November 2020–July 2021), we found two outliers (**Figure 1B**). In the cohort conceived in January 2021 and exposed in months 6–7 of pregnancy, the odds difference was 16.1 (14% increase, 95% CI: [5%, 24%]). The odds of preterm birth in this cohort were 112.3 per 1000. The May 2021 cohort, exposed in months 2–3, had an odds difference of 12.3 (11% increase, 95% CI: [1%, 22%]), and the odds of preterm birth in this cohort were 110.1 per 1000.

## Discussion

The 2021 PNW Heat Dome caused historic high temperatures in an otherwise temperate region unprepared for heat. Our analysis showed that the Heat Dome increased odds of preterm birth in conception cohorts exposed in months 2-3 and 6-7 of pregnancy by 11% and 14%, respectively.

Our results align with prior literature on heat and preterm birth, which includes studies largely conducted in frequently-exposed populations.^9–11^ A 2020 meta-analysis of six studies, mostly assessing short-term heat exposure in the week preceding delivery, found that heatwave exposure (two or more days above a prespecified threshold) was associated with 16% (95% CI: 10-23%) higher odds of preterm birth.^11^A subsequent 2024 meta-analysis including additional studies found 26% higher odds (95% CI: 8% - 47%) of preterm birth following heatwave exposure four weeks before delivery.^10^ We found increased odds of preterm birth using a different method, in a historically unexposed study population, and considering exposure throughout pregnancy.

Prior studies report heat-preterm birth associations across all trimesters.^17–20^ We examined exposure monthly rather than trimesterly, which is more granular than previous literature. The literature suggests that heat may increase preterm birth risk throughout gestation, potentially through different mechanisms at different stages of pregnancy. For example, heat may impair placental development in early pregnancy,^21,22,23^ increase cardiovascular strain in both early- and mid-pregnancy,^24,25^ and increase uterine contractility in later pregnancy.^26^ Heat exposure may also induce conditions like gestational hypertension and preeclampsia, which in turn increase preterm birth risk.^21,26–28^

Most previous studies consider non-emergency variation in heat exposure, while we studied an extreme anomalous event. During the PNW Heat Dome, pregnant people may have been exposed to higher temperatures, experienced stress in addition to heat, or changed their behavior to protect themselves from heat differently compared to non-emergency situations, potentially influencing our susceptible exposure window results.

Our quasi-experimental design and synthetic control time series allowed us to minimize bias from observed, unobserved, and time-varying confounding. However, we assessed exposure based only on birthing person residence in Oregon or Washington at delivery. We did not capture variation in heat exposure by housing quality, greenspace access, urban heat island effect, outdoor work or travel, or air conditioning access.^29–31^ We did not examine at-risk subgroups (e.g., persons with gestational hypertension or diabetes), who may be more susceptible to preterm birth from heat exposure.^9–11^

Historical and present-day racism and segregation determining neighborhood characteristics like greenspace and shade, as well as psychosocial stress, economic opportunity, and healthcare access means extreme heat exposure and susceptibility are more common in racialized people, especially Black people.^32^ Increasing heat exposure has the potential to exacerbate existing preterm birth disparities,^33^ and future studies should incorporate environmental justice analyses. They should also consider additional perinatal outcomes such as stillbirth, which heat exposure may impact via similar mechanisms to preterm birth.^9–11,26^

Heatwaves like the PNW Heat Dome will occur more frequently with climate change.^4^ Further studies should test interventions to mitigate heat impacts on perinatal health, following the lead of community-based organizations.^34^ Potential interventions include improved communication about heat risks during pregnancy, expanded access to prenatal emergency care, stronger worker protections for pregnant people, and municipality-scale interventions to reduce exposure, such as landlord requirements for cooling,^35^ heat response plans,^36^ and greening initiatives.^37^

## Data Availability

This study is based on restricted vital statistics data, which is available by application from the CDC: https://www.cdc.gov/nchs/nvss/nvss-restricted-data.htm.

## Funding

Canadian Doctoral Foreign Study Award (HM)

NIHLBI T32 HL140290 (NMF)

NIEHS P30ES007033 (JAC)

NICHD R01HD103736 (RC, TB, AS, AG, JAC)

## Notes

### Competing Interest Statement

The authors have declared no competing interest.

### Funding Statement

This work is supported by the following funding: Canadian Doctoral Foreign Study Award (HM),
NIHLBI T32 HL140290 (NMF),
NIEHS P30ES007033 (JAC),
NICHD R01HD103736 (RC, TB, AS, AG, JAC).

### Author Declarations

The UC Irvine Committee for the Protection of Human Subjects approved this study (protocol # 20195444).

